# Study protocol for a multi-centre stepped-wedge cluster randomised trial to explore the usability and outcomes among young people living with HIV in Kiambu and Kirinyaga counties of Kenya, using an online health portal

**DOI:** 10.1101/2023.10.04.23296464

**Authors:** Eric Nturibi, Jared Mecha, Elizabeth Kubo, Albert Orwa, Florence Kaara, Faith Musau, Christine Wamuyu, Justus Kilonzi, Randeep Gill, Sanne Roels

## Abstract

**Introduction:** While the incidence of HIV declines in most age groups globally, it continues to rise among adolescents and young adults who also experience a higher rate of HIV-related deaths. These tech-savvy individuals might benefit from an online patient portal aimed at increasing awareness, building skills, and promoting patient activation – the willingness and capacity to independently manage their health. However, the impact of such portals on young HIV patients in Kenya remains uncertain.

**Methods and analysis:** HIV patients aged 15-24 with smartphone access will participate in a 12-month stepped wedge cluster randomized trial. The main focus will be the portal’s effect on patient activation, with secondary outcomes being self-reported adherence, viral suppression, and social involvement. The study also aims to understand the portal’s development, implementation, functionality, usability, and cost. From 47 antiretroviral therapy (ART) sites with electronic medical recording systems, 16 clusters of 30 participants each will be formed. These clusters, over 12 months, will be randomized into 3 intervention sequences every 3 months. Baseline measurements, covering patient activation, adherence, viral load, and social engagement, will be recorded over two weeks, with subsequent checks at 3, 6, and 12 months. Data will be analyzed using generalised linear mixed models.

**Ethics and dissemination:** The protocol has received approval from the AMREF Health Africa Ethics & Scientific Review Committee (ESRC) and local governments. Findings will be shared through stakeholder forums, conferences, publications, and media.

**Author Summary:** *What is the issue?:* HIV infections among teenagers and young adults are increasing, particularly in Kenya. Given their technological inclination, an online portal might help them manage their health. The effectiveness of this approach remains uncertain.

*What are our actions?:* We are running a year-long study in Kenya with HIV-positive individuals aged 15 to 24 who have smartphones. The goal is to see if the portal aids their health management. We’re assessing health readiness, medication adherence, HIV control, and social connection. The study involves groups from 47 HIV treatment centers, with new groups engaging with the portal every three months.

*What are our assessment methods?:* We will use statistical methods to gauge the portal’s impact on health management.

*Is the study approved?:* Yes. It has endorsements from AMREF Health Africa and local governments. Results will be shared with the community, at conferences, in scientific journals, and in the media.

## Introduction

Of the 37.7 million persons living with HIV (PLHIV) worldwide in 2020, 25.3 million were resident in sub-Saharan Africa (SSA), where 460,000 of the 680,000 global AIDS related deaths (ARDS) also occurred (UNAIDS, 2021). In 2020, 400,000 adolescents and young persons (AYP) were newly infected with HIV, of whom 150,000 were aged 10 to 19(1). Globally, whereas new HIV infections are declining for other age groups, they are increasing in the adolescent age group (10-19yrs), who also contribute disproportionately to ARDs(2). In 2020, there were an estimated 1.4 million PLHIV in Kenya including 160,000 young persons (ages 15-24). Of the estimated 33,000 new infections, 35% occurred in young persons (ages 15-24)(3). With 59.24 million connections as at January 2021, Kenya had a mobile penetration of 108.9% and an internet penetration of 40% (21.75 million), including 11 million social media users(4). According to a report by the United Nations, young people in low and middle income countries (LMICs) are nearly three times more likely to be using the internet than the general public(5). A recent review of digital health solutions (DHS) for engaging PLHIV decried the lack of published interventions targeting the more than 4 million young people aged 15-24 living with HIV globally, despite their identification as a “critical group experiencing multiple barriers to engagement in care.”(6). Patient health portals are therefore particularly attractive in this vulnerable population who are likely to welcome digital health solutions more readily than other age groups.

The functionalities proposed in the digital health platform, myCareHub™, have been carefully designed to incorporate mechanisms identified as critical for outcome achievement (fig 1) (7) and are based on the COM-B framework (fig 2) that links behaviour to capability, opportunity, and motivation (8).

**Figure 1:**
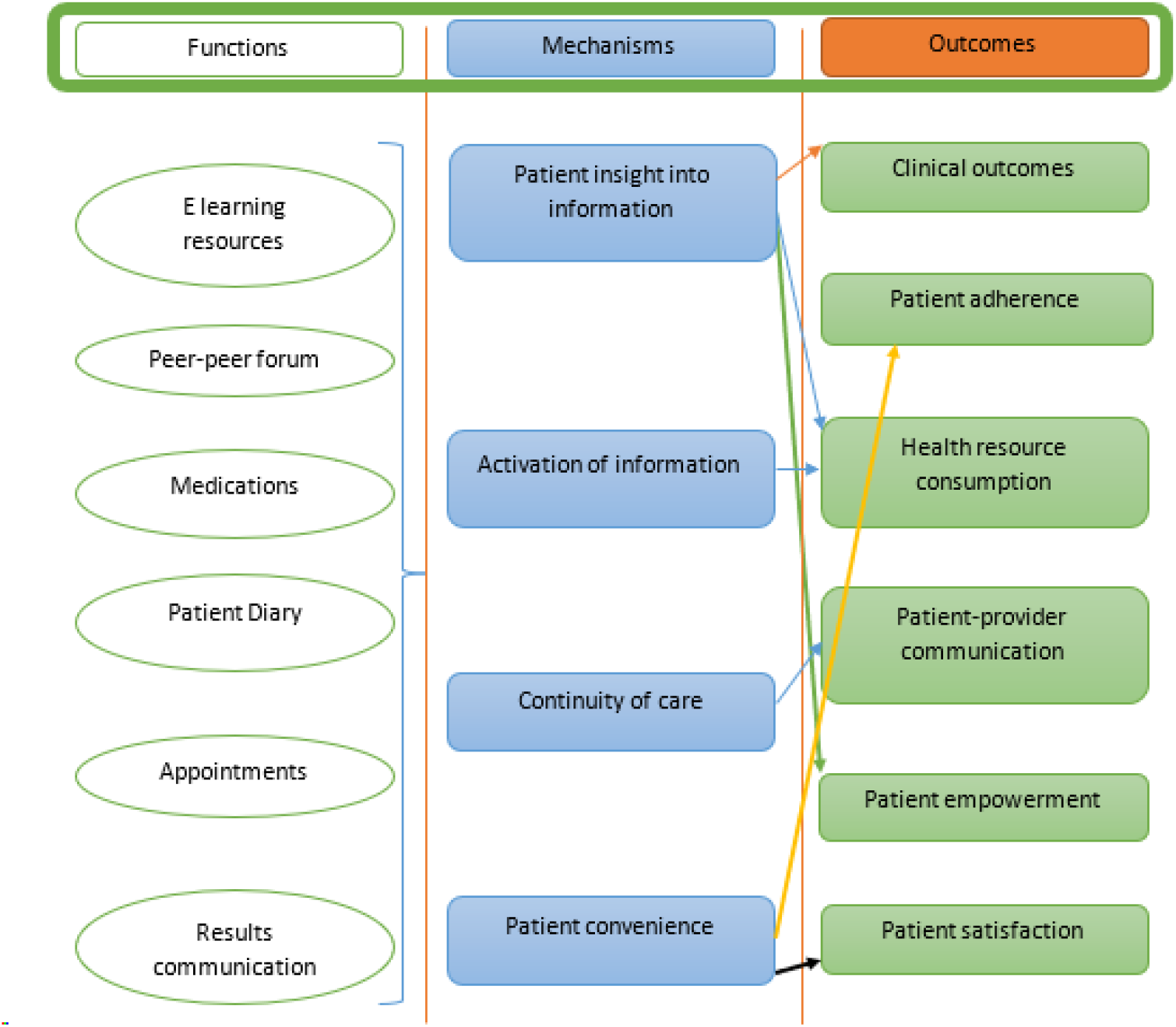
Mechanisms of patient activation (adapted from Otte-Trojel, de Bont (7))

**Figure 2:**
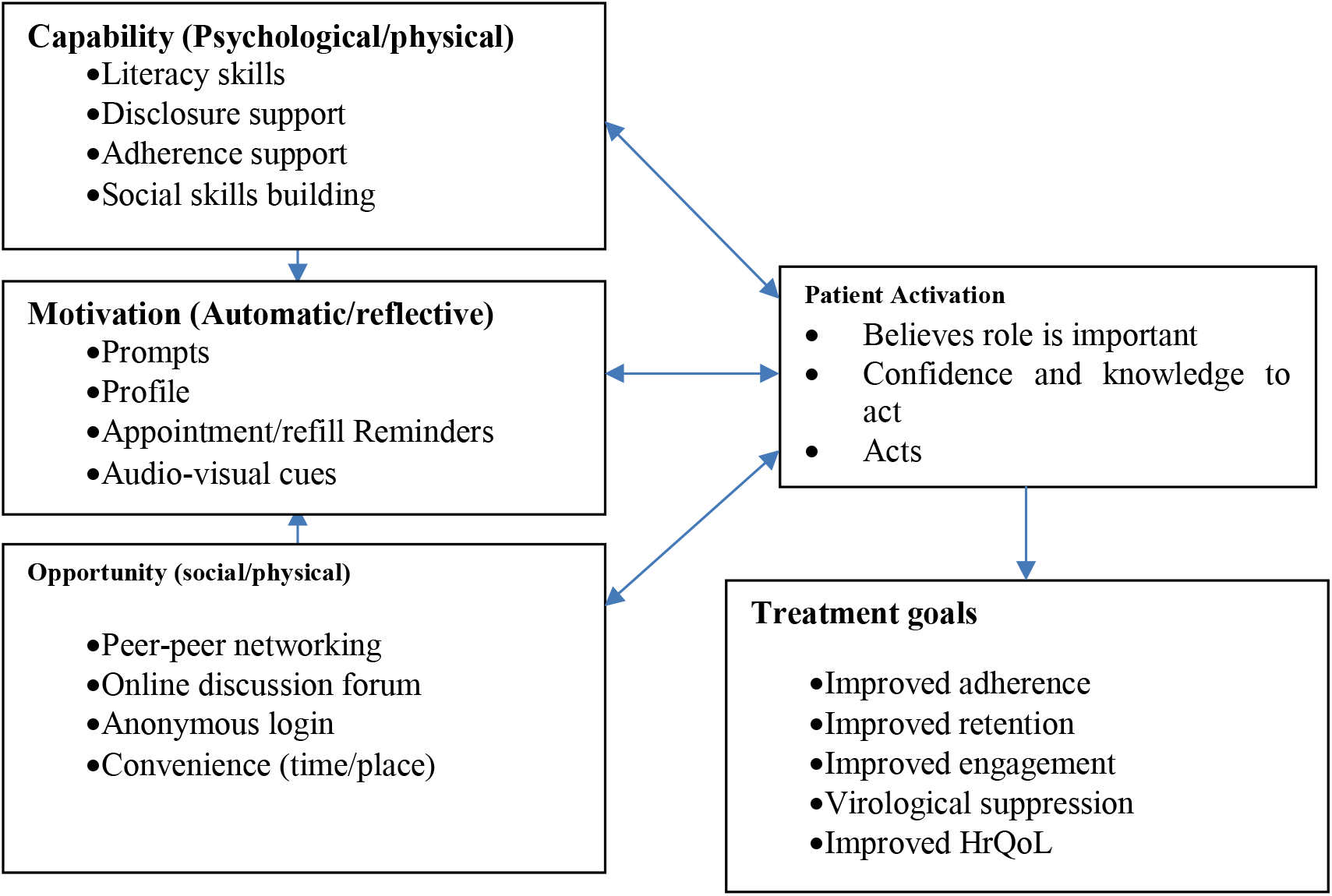
COM-B theoretical framework for behaviour change.

The design was heavily influenced by the views and preferences of the intended users (unpublished data) with inputs from an experienced team of AYP resource persons. **myCareHub™** will be accessible on internet enabled mobile phones through a dedicated password to ensure security and confidentiality of data. It is designed to offer a wide array of services to healthcare providers (HCPs) and patients (fig 3).

**Figure 3:**
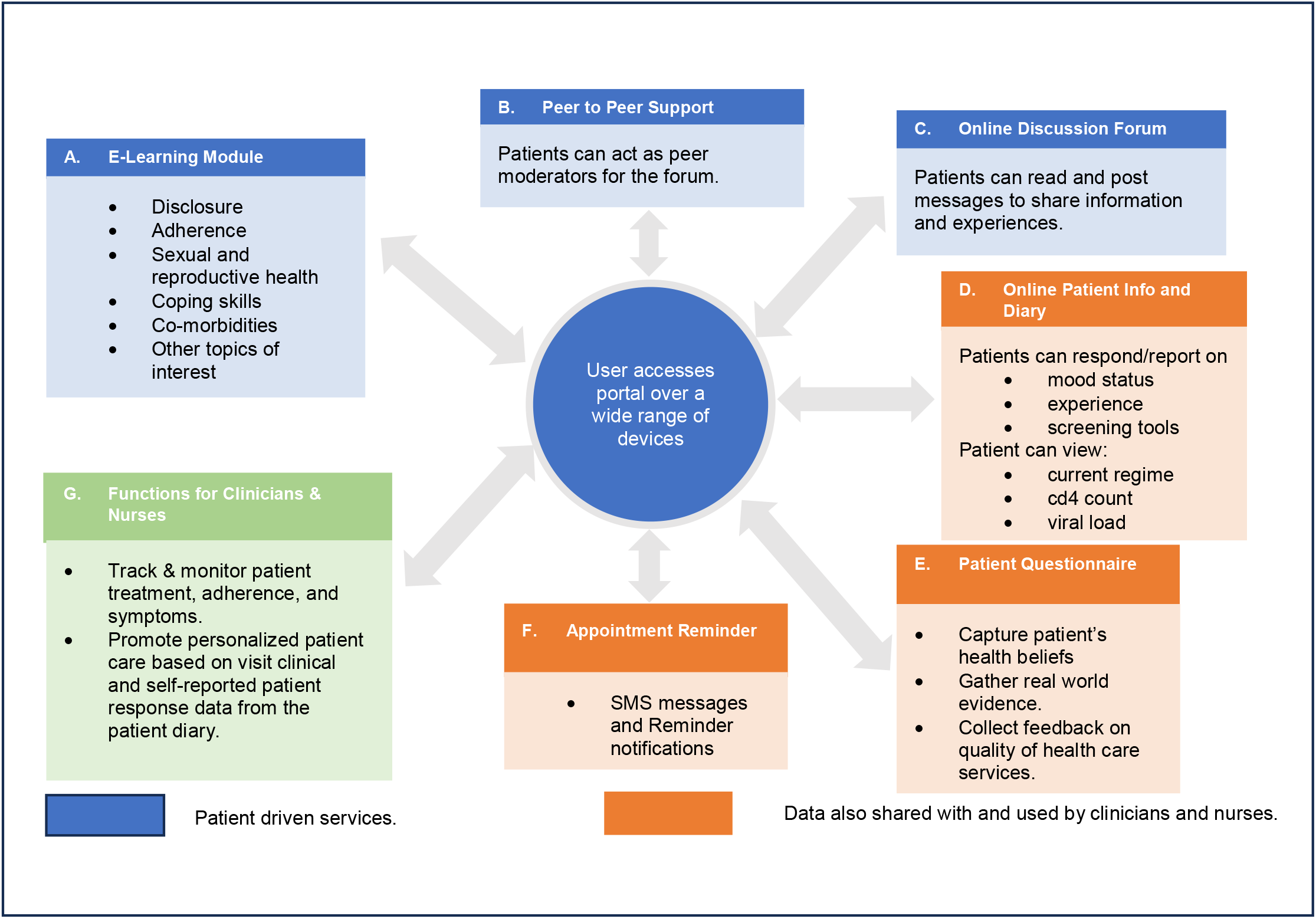
Diagrammatic representation of myCareHub™ and functions available to use.

This study seeks to establish whether an online patient health portal, myCareHub™ improves AYP activation and consequent engagement in HIV care and treatment. Further, in line with World Health Organization (WHO) guidelines for mobile health evidence reporting and assessment (mERA), the study will explore quality, fidelity, and coverage of the platform, as well as key program indicators (fig 4)(9); features that are often overlooked in published studies.

**Figure 4:**
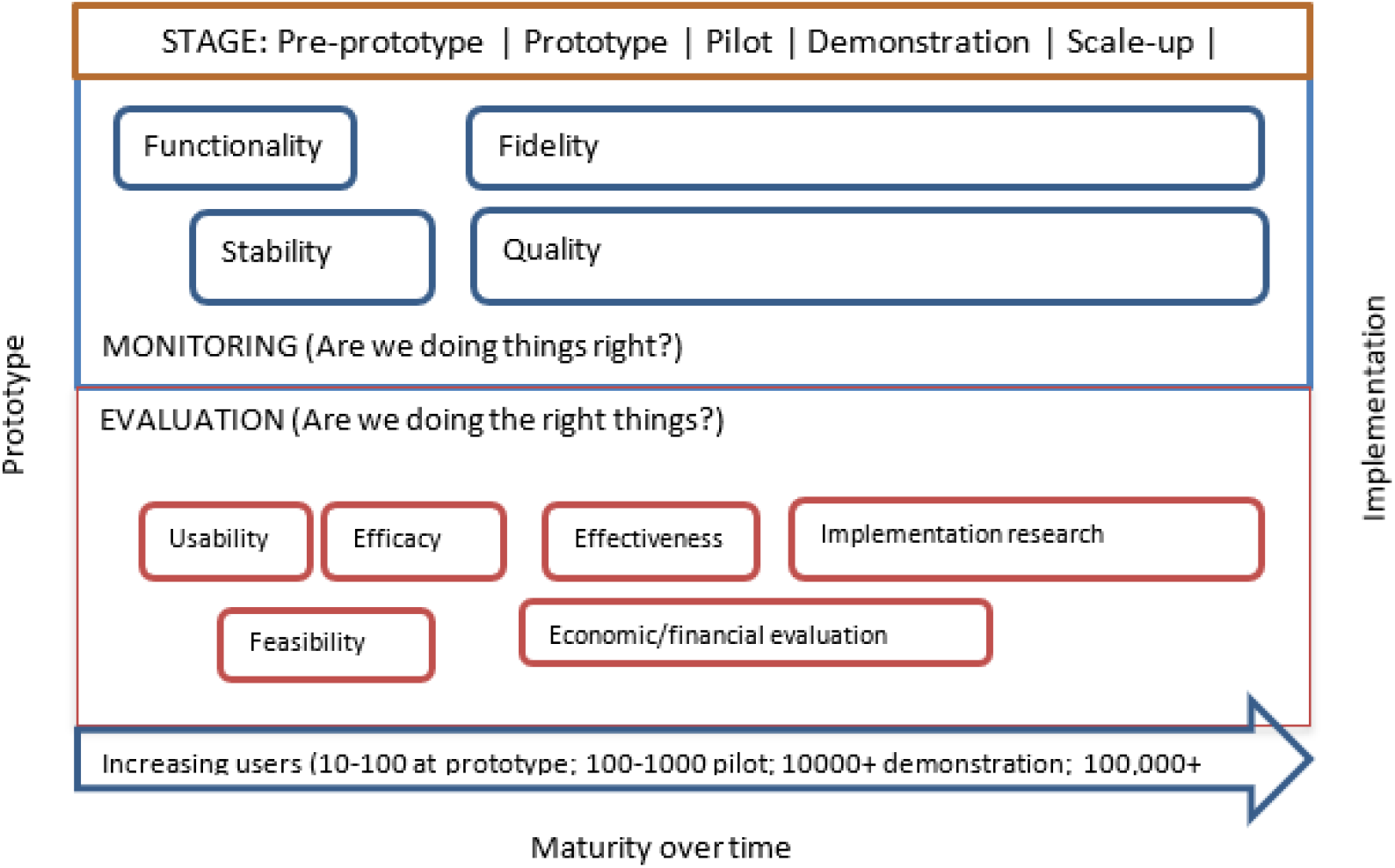
Intervention life cycle of a digital intervention.

The findings of this study will inform HIV programming for young persons and other vulnerable populations, shift health policy on digital health solutions and contribute substantially to the elimination of AIDS as a global threat by 2030.

## Methods and Analysis

### Setting

The University of Nairobi – Central Kenya Response: Integration, Strengthening and Sustainability Plus (CRISS Plus) project is a 5-year PEPFAR-funded HIV prevention, care & treatment project in partnership with Kiambu & Kirinyaga county governments. The project currently supports HIV related activities at 78 public facilities. Forty-seven (47) facilities have an electronic medical records (EMR) system. The 47 EMR sites will be clustered based on geographical proximity loosely coinciding with sub county territorial assignment of the facilities and covering 15 out of the 17 supported sub-counties. The largest treatment site will constitute the 16^th^ cluster on account of the large number of adolescent and young people enrolled in its antiretroviral treatment (ART) program.

At study inception, the program provided ART for 38,079 PLHIV, of whom 3,459 (9%) were AYP. The target population for the study will be AYP (ages 15-24 years) enrolled in supported clinics. Eighty-nine per cent (89%) of the target population are enrolled in the 47 EMR sites.

### Study design

We plan to conduct a multicentre stepped wedge cluster randomised trial to assess whether a bespoke patient health portal, myCareHub™, can improve patient activation and clinical outcomes among consenting individuals aged 15 to 24 living with HIV and attending the ART clinic at one of 47 EMR-enabled treatment centres (table 1). Caregiver consent will be obtained for participants under the age of 18.

**Table 1:** PICOT Framework.

| *Table 1: PICOT Framework* |  |
| --- | --- |
| Population | Adolescents and young persons (15-24 years old) attending an EMR-enabled Anti-Retroviral Treatment (ART) site and with ready access to a smartphone |
| Intervention | Making a patient health portal with several functionalities available |
| Comparator | Standard of care patient engagement strategies |
| Outcome | Patient activation as the primary outcome. Secondary outcomes: viral load, social engagement measures, adherence, and portal use features |
| Time | 15 months total, the first three being control, with 3-month intervals between sequences, and measurements taken over a 2-week period. Once all clusters cross over from control to intervention, an observation period of 6 months will follow. |

In total, a minimum sample of 480 adolescents and young persons will be recruited into 16 clusters using a stepwise design (table 2) with intervals of three months between successive steps.

**Table 2:**
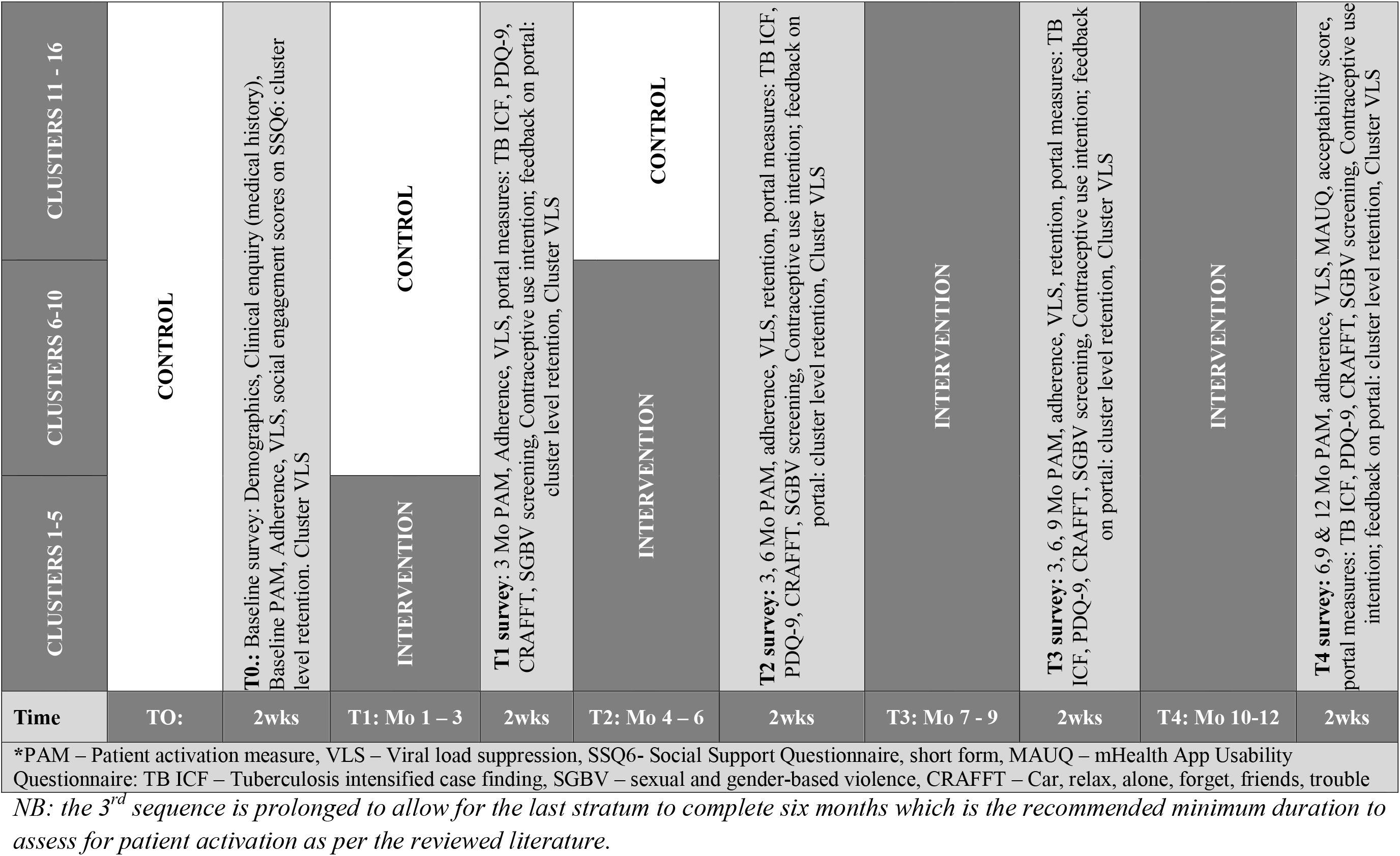
Study design: Cluster randomized, stepped wedge design with 16 clusters and 3 sequences.

Stratification of clusters will be conducted based on size, sex/gender mix and viral load suppression (table 3) to come up with comparable blocks of clusters prior to randomisation (table 4).

**Table 3:** Sub-counties by key attributes.

| Sub-county | Number of AYPs | Size Strata | *VLS 15-19 | VLS 20-24 | Cohort retention | Males (%) |
| --- | --- | --- | --- | --- | --- | --- |
| Thika town | 632 | Large | 80% | 90% | 89% | 41% |
| Ruiru | 330 |  | 84% | 89% | 68% | 25% |
| Kiambu town | 205 | Medium | 86% | 89% | 100% | 32% |
| K Central | 176 |  | 86% | 90% | 84% | 55% |
| Gatundu South | 166 |  | 61% | 90% | 50% | 40% |
| Juja | 146 |  | 91% | 82% | 67% | 35% |
| Kiambaa | 123 |  | 87% | 91% | 100% | 27% |
| Kirinyaga South | 116 |  | 91% | 87% | 88% | 53% |
| Githunguri | 113 |  | 85% | 89% | 100% | 27% |
| Kabete | 110 |  | 94% | 94% | 100% | 23% |
| Gatundu North | 107 |  | 92% | 89% | 100% | 40% |
| Limuru | 103 |  | 90% | 100% | 100% | 35% |
| Kirinyaga West | 88 | Small | 81% | 92% | 74% | 37% |
| Kikuyu | 73 |  | 100% | 90% | 100% | 16% |
| Kirinyaga East | 58 |  | 100% | 86% | 87% | 57% |
| Lari | 51 |  | 100% | 91% | 100% | 23% |
| Kirinyaga North | 50 |  | 60% | 68% | 88% | 48% |
| *VLS – viral load suppression |  |  |  |  |  |  |

**Table 4:** Randomization of clusters into sequences.

| S/N<br>o. | Cluster | 15-19<br>yrs. | 20-24<br>yrs. | Total<br>AYP | Strata<br>Size | Rando<br>m<br>numbe<br>r | Se<br>q | No. of<br>AYP<br>(n=48<br>0) |
| --- | --- | --- | --- | --- | --- | --- | --- | --- |
| 1 | Juja | 48 | 98 | 146 | Medium | 0.91499 | 1 | 186 |
| 2 | Kiambaa | 30 | 80 | 110 | Medium | 0.842108 | 1 |  |
| 3 | Kirinyaga<br>Central | 76 | 83 | 159 | Medium | 0.637954 | 1 |  |
| 4 | Kabete | 19 | 75 | 94 | Small | 0.969301 | 1 |  |
| 5 | Kirinyaga South | 27 | 48 | 75 | Small | 0.832712 | 1 |  |
| 6 | Ruiru | 77 | 253 | 330 | Large | 0.725233 | 1 |  |
| 7 | Gatundu South | 75 | 91 | 166 | Medium | 0.555849 | 2 | 170 |
| 8 | Gatundu North | 52 | 55 | 107 | Medium | 0.546696 | 2 |  |
| 9 | Limuru | 41 | 62 | 103 | Medium | 0.541296 | 2 |  |
| 10 | Lari | 12 | 23 | 35 | Small | 0.701539 | 2 |  |
| 11 | Kirinyaga East | 29 | 26 | 55 | Small | 0.567749 | 2 |  |
| 12 | Thika Level 5 | 172 | 196 | 368 | Large | 0.226127 | 2 |  |
| 13 | Githunguri | 45 | 56 | 101 | Medium | 0.458081 | 3 | 124 |
| 14 | Thika Town | 102 | 135 | 237 | Medium | 0.379402 | 3 |  |
| 15 | Kiambu<br>Township | 87 | 112 | 199 | Medium | 0.062999 | 3 |  |
| 16 | Kirinyaga West | 29 | 42 | 71 | Small | 0.514886 | 3 |  |

Clusters within each comparable block (stratum) will be randomly assigned to one of the three sequences for time of crossover from control to intervention using a computer-generated list of random numbers. Clusters will receive the intervention in groups of five and six. There will be a total of 3 intervention sequences – the third extending 6 months. Each cluster will begin in the control period and eventually receive the intervention, with the order of crossover being randomly determined. An interval of two weeks between intervention sequences will be incorporated to allow for measurements to be taken and the intervention to be introduced to new clusters.

### Justification of stepped wedge design

The stepped wedge design was chosen as individual randomization into control and the patient portal within a facility would inevitably lead to contamination as the control group would inadvertently be exposed to the intervention. The staggered approach to implementation also allows for enhancements to the portal based on lessons learned from preceding clusters.

### Study population

In our study, the intervention will be targeted at clusters, where a cluster is defined as a group of EMR-capable facilities in geographical proximity. Outcome parameters will be determined at patient level. Two key strategies will be employed to recruit participants into the study. A line listing of all AYP in the participating EMR sites will be done from the central EMR database and random number sequences used to select eligible participants per facility with allowance made for refusals. Selected participants will then be invited for an open day activity during which recruitment will be done after obtaining informed consent. Weekly appointment diaries will also be used to line list AYP due for their clinic visit in the coming week with a random number sequence generated and applied to select eligible participants. These strategies will be undertaken repeatedly until the required sample size is obtained. Measurements will be taken on the same cohort and by the same observer at the end of each successive step when a cluster moves from control to the active intervention (table 2).

### Intervention

myCareHub™ will be available on google play-store and iOS app-store. The portal has been developed by the study team with significant input from end users and is designed to offer several functionalities critical for catalysing the behaviour change process as per the COM-B framework (figs 2&3). The portal will comprise two mobile based applications: a patient portal and a health care provider (HCP) portal. Features available to patients will include e-learning resources in text, audio, videos and other media, focussing on HIV treatment literacy, disclosure support, adherence support, sexual and reproductive health (SRH) and coping skills; online communities for peer-to-peer networking and learning; an online patient diary to support self-monitoring by enabling patients to view their medical data from the facility based EMR and to record and share their experiences with HCPs; online based web forms and survey tools to capture patient reported outcomes which will trigger a service request on the HCP portal so that care can be personalised and offered without delay; in app and short message service (sms) notifications for appointments and other reminders; and a mechanism for appointment rescheduling (to be developed in the second phase). The HCP portal will enable providers to track and monitor patient treatment, adherence, and symptoms, and to provide personalised care based on response data from the patient diary. As the apps are linked and synchronised with the facility EMR, HCPs can capture and access longitudinal patient level visit data and other medical information such as laboratory investigation reports. On a quarterly basis, open forums will be conducted with portal users - HCPs and patients, to ensure they can access and use all the available features and to capture qualitative end-user insights.

### Outcome parameters

Outcome parameters will be measured in individuals at pre-specified time points, baseline, every 3 months, and end-line (table 5), using digitised case report forms.

**Table 5:**
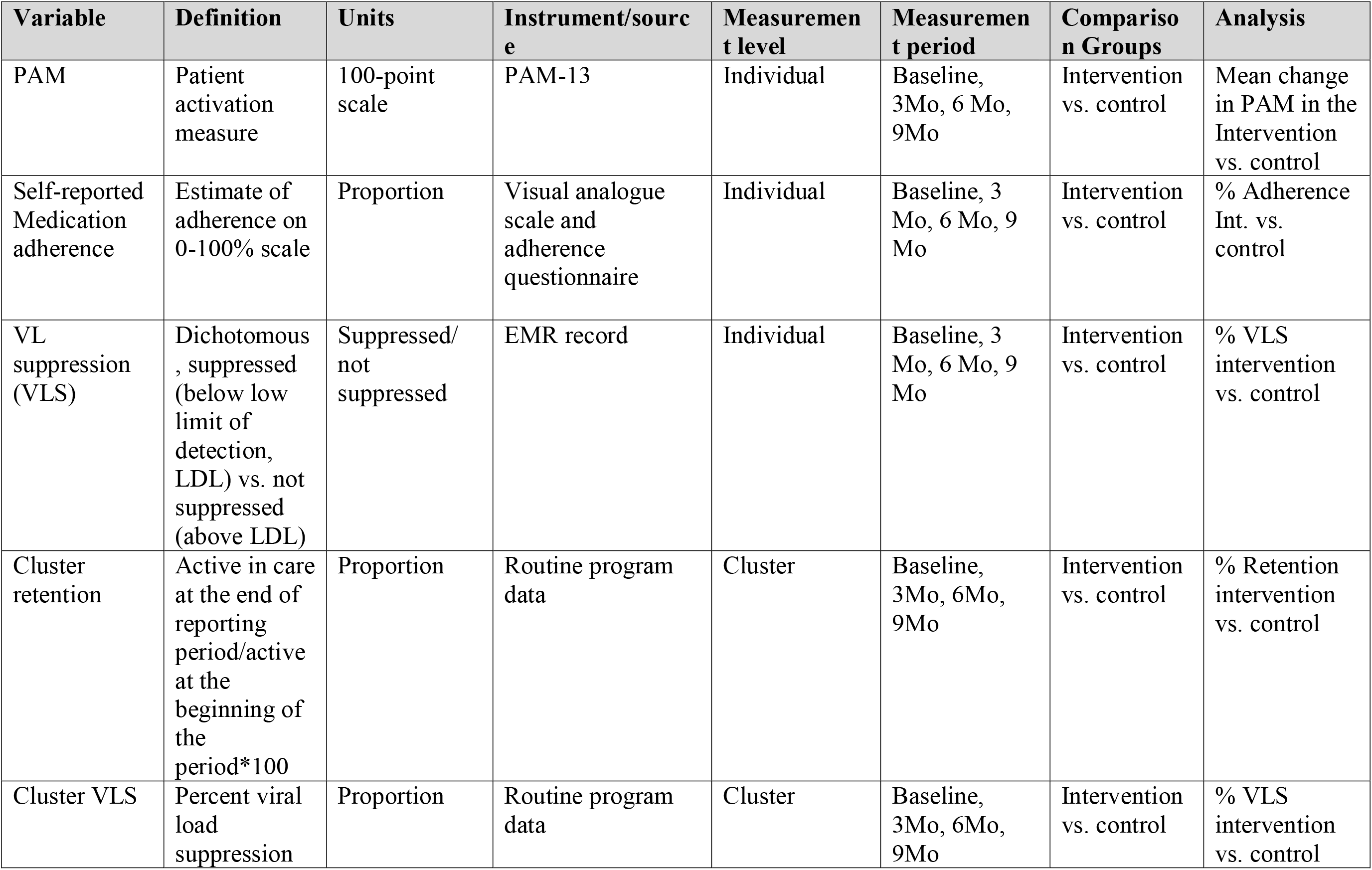
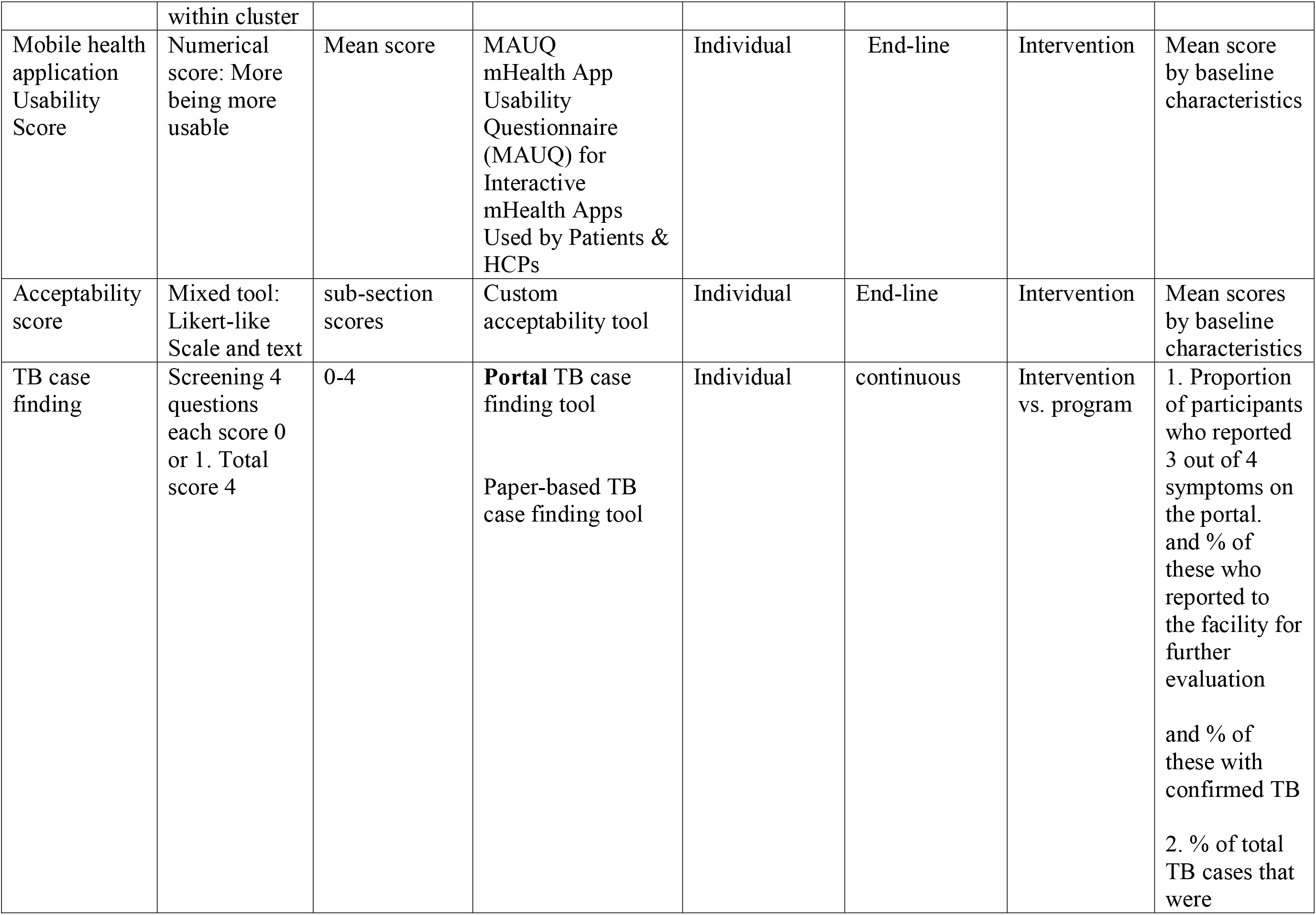
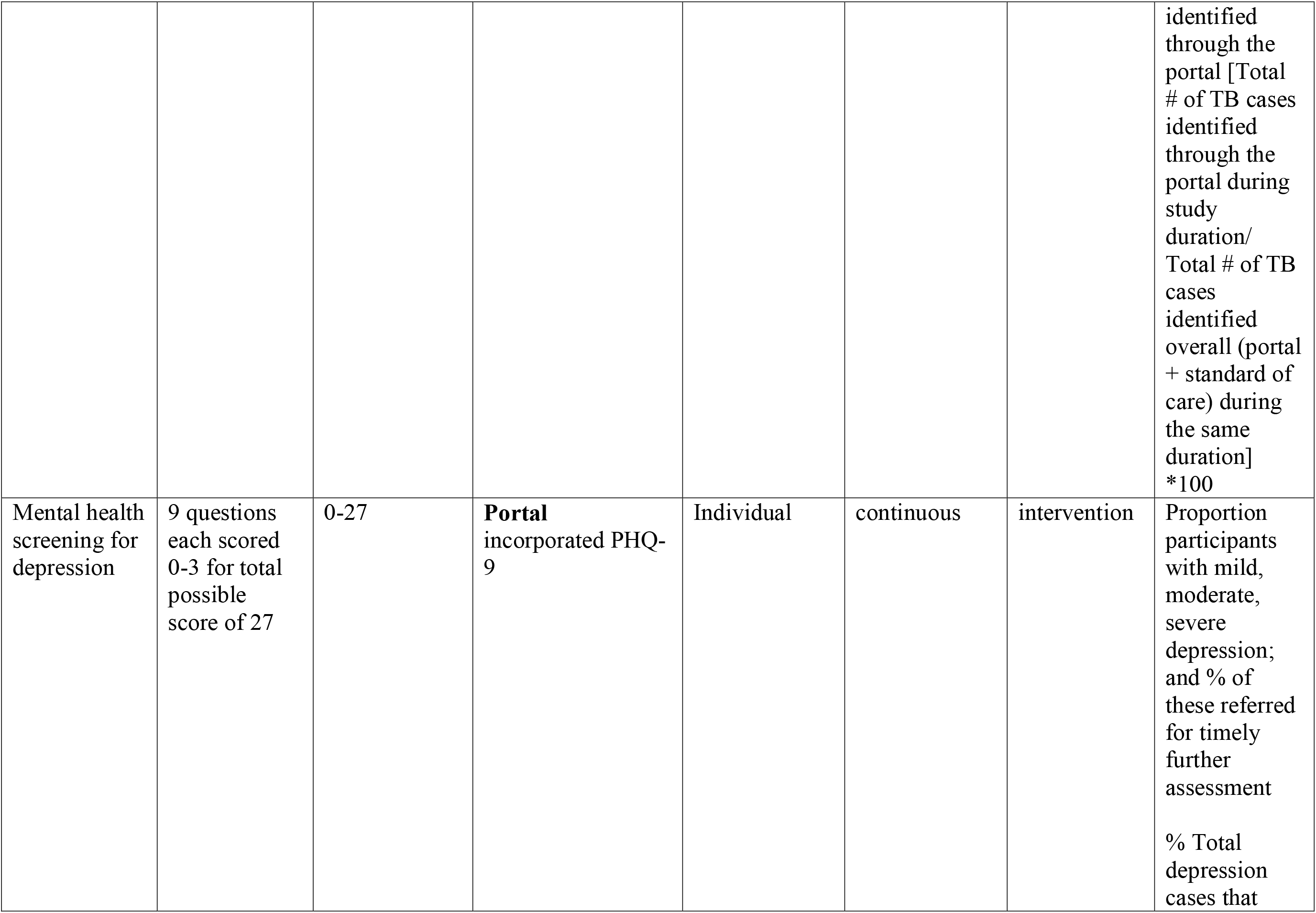
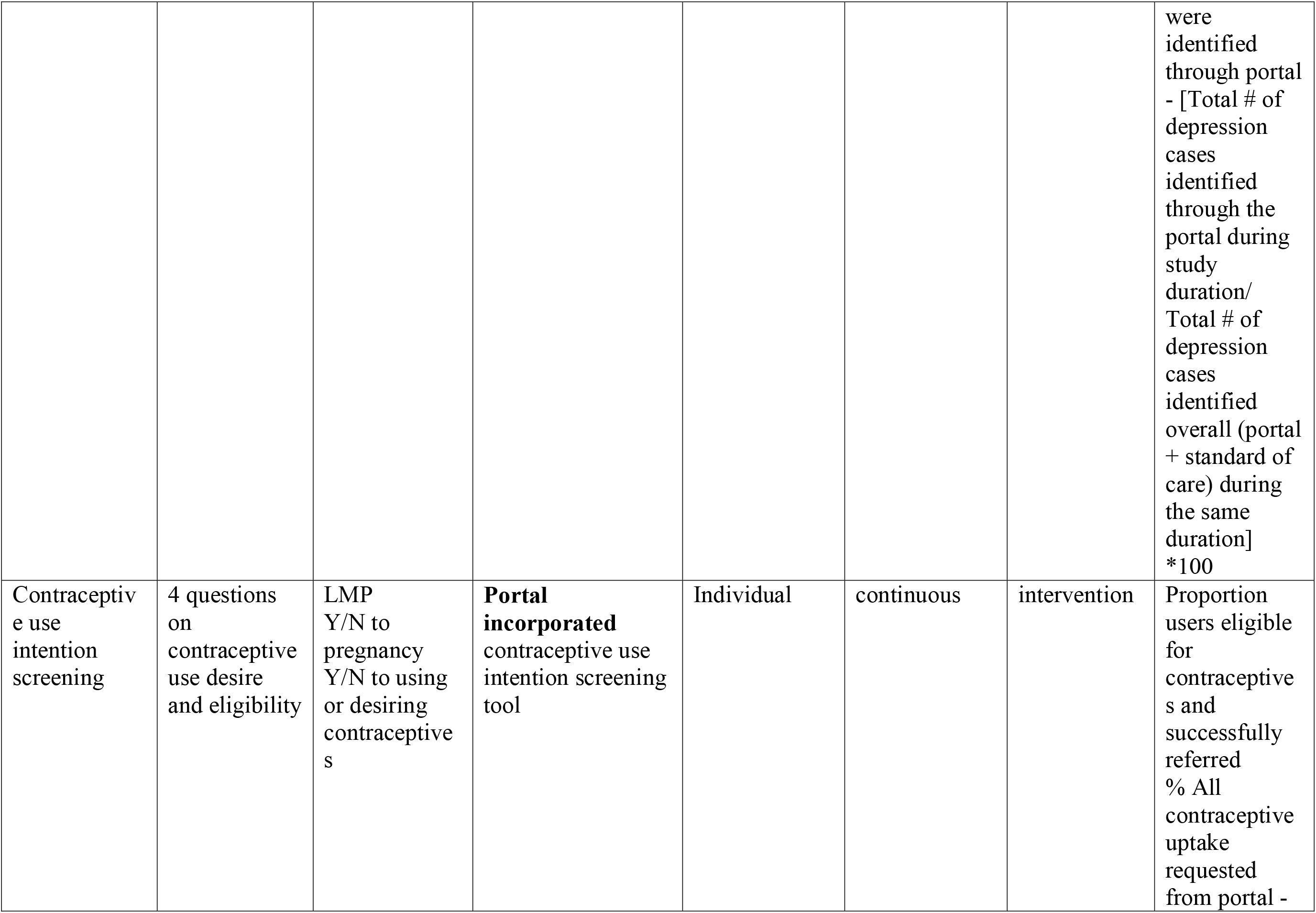
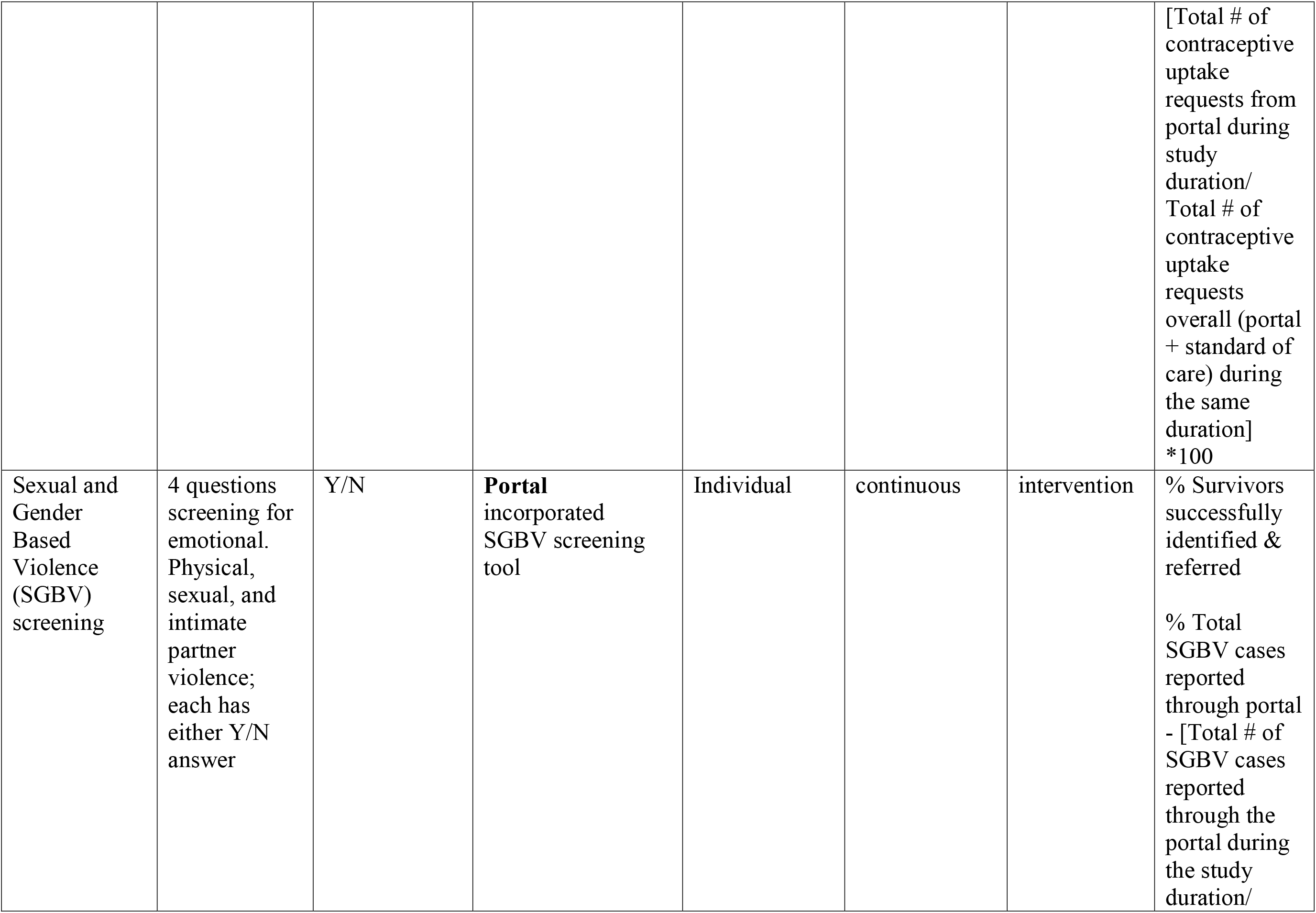
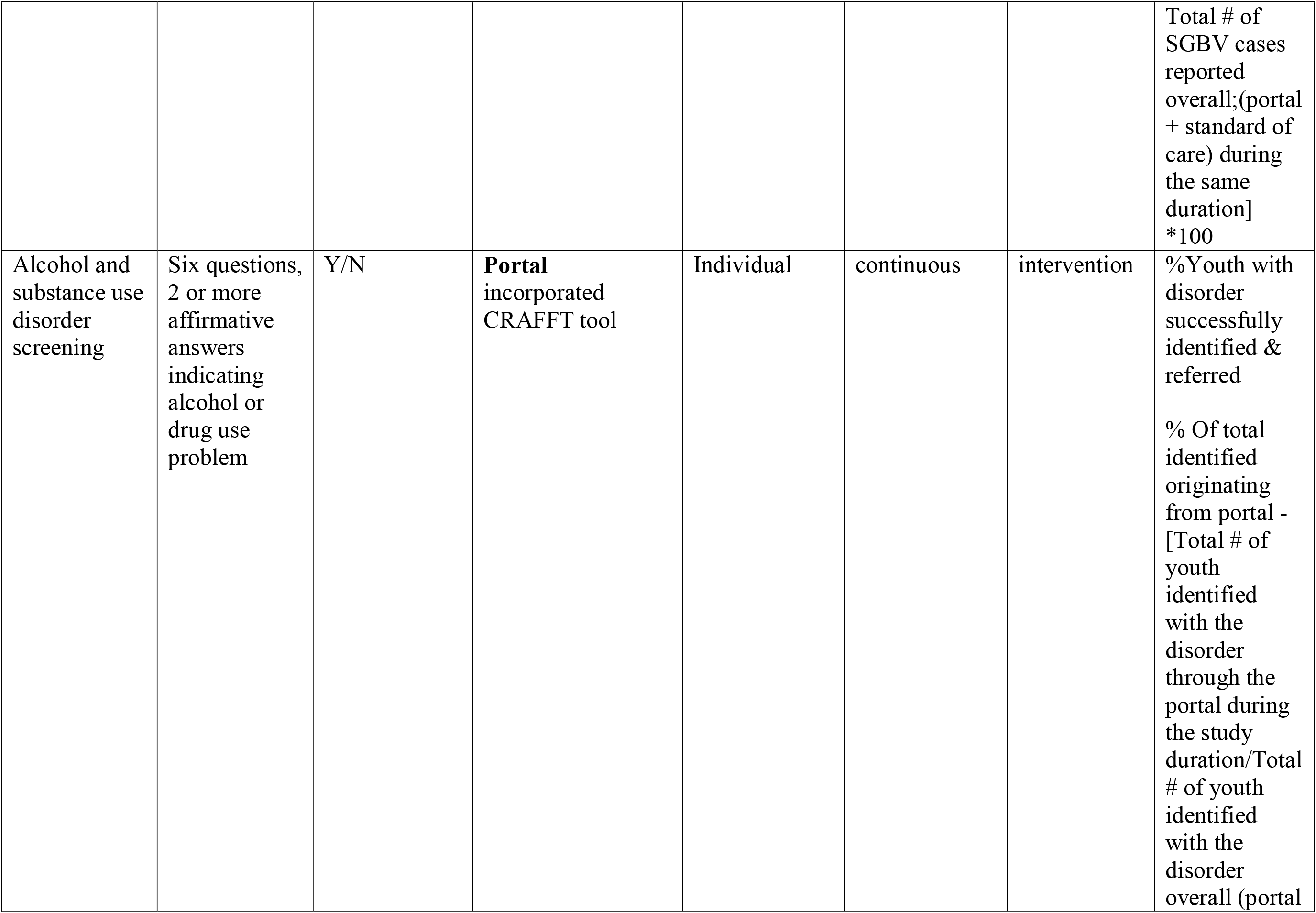
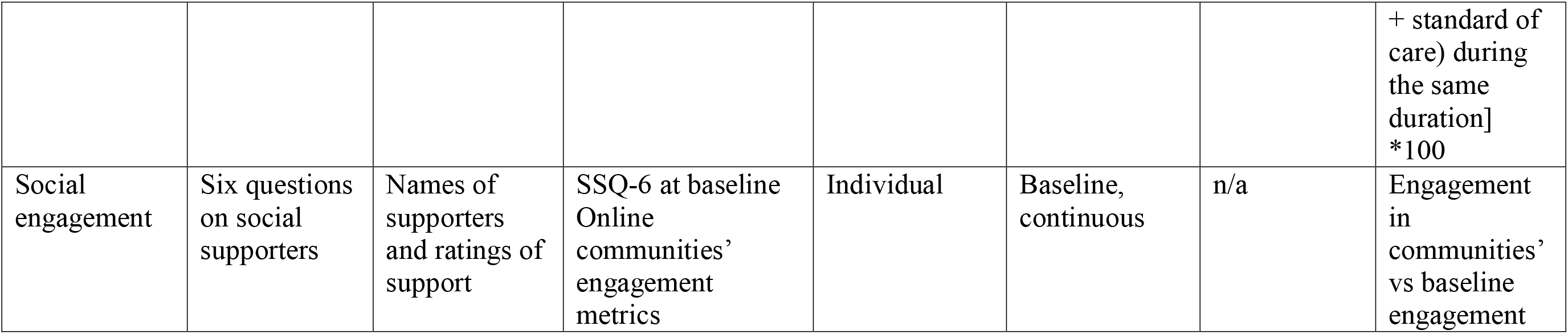
Outcome variables.

The primary outcome will be change in patient activation measure (PAM) at 6 and 9 months. Secondary outcomes will include self-reported adherence (months 3,6 &9) and latest viral load.

### Portal-based measures

Portal-based measures will include portal functionality, quality, fidelity, and an estimate of total cost in setting up the portal. Four key elements of the patient diary – screening for Tuberculosis (TB) and sexual and gender-based violence (SGBV), contraceptive needs assessment and screening for alcohol and substance use disorders will be monitored, and the time taken for HCPs to resolve service requests recorded and compared to usual non-portal based clinical practice. An analytical dashboard will be developed to provide real time updates on portal engagement metrics.

### Evaluating Usability

Participation in online communities will be tracked and compared with baseline social engagement score (SES).

During the study, focus group discussions (FGDs) will be used to elicit insights into the client experience of the portal that will go towards future enhancements.

A mobile health app usability survey (MAUQ - patient and health care provider) and an acceptability survey will be administered at the end of the intervention period.

### Statistical methods

Baseline participant characteristics i.e., sociodemographic (age, sex, occupation) and clinical information (duration on ART, presence of treatment buddy and last viral load) will be described by cluster.

The primary outcome will be analysed as mean PAM/ change in PAM score between intervention and control groups at 6 and 9 months by age category (15-19 vs. 20-24), sex (male vs. female) and duration on ART (<12 months and >12 months). PAM total scores of 0 and 100 will be considered as missing data and PAM changes will be analysed over time and across PAM levels as per the developer’s recommendations.

Statistical analysis of the primary and secondary endpoints will be conducted in either R or SAS using generalised linear mixed models which allow for incorporation of correlations due to repeated measures, cluster effects and potential time effects. Sensitivity analyses, including use of causal inference methods, will be conducted to assess the effect of differential patient and/or cluster characteristics over time and/or attrition on the comparison between intervention periods.

Thematic analysis will be conducted on qualitative insights from FGDs, and the operational and other costs related to running the portal tracked to facilitate cost analysis.

### Sample size

The primary outcome will be a change in patient activation measure (PAM) (Hibbard JH, et al, 2004). Lin and co-authors found an average effect size of 0.33 in a meta-analysis on studies involving patients with chronic disease reporting change in PAM (Lin et al., 2020). Also, a difference of 4 points on the PAM scale is considered clinically important and Marshal et al were able to demonstrate that a 5-point difference was significantly associated with higher odds of CD4 count>200, retention, and viral load suppression (J. H. Hibbard & Tusler, 2007; Rebecca Marshall et al., 2013). Following the definitions as outlined in Hooper et al., 2006, assuming an intra-cluster correlation of 0.02 and individual autocorrelation of 0.8, and only low “time by cluster” effects impacting the PAM scores, the study will require a minimum sample size of 27 per cluster to demonstrate a change of 5 point (green lines in figure 5) in the activation score between the two groups, assuming the same standard deviation as in Marshall, Beach (10) with a power of at least 90% at an alpha of 0.05% (11).

**Figure 5:**
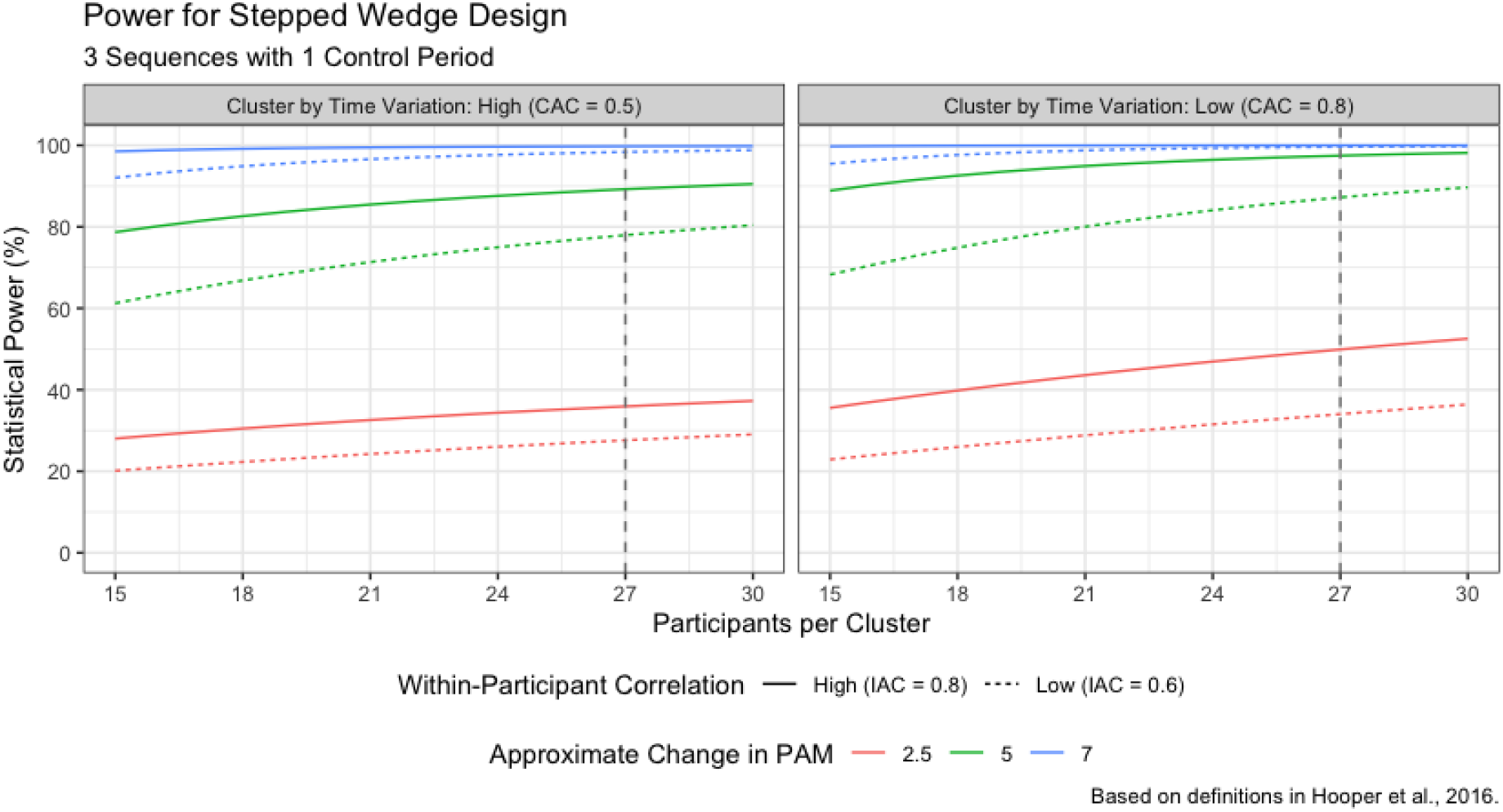
Sample size estimation for stepped wedge design.

It is worth noting that the power to demonstrate a difference will be affected substantially by the cluster by time effect. Hemming et al. provided minimal guidance – but did mention a range on that correlation between 0.3 (large effect) to 0.9 (small effect). They strongly suggest a sensitivity analysis on these assumptions, which is provided in Figure 5. Focusing on the green lines, in the presence of high intra-participant variability but small time by cluster effects, approximately 90% power can be attained with 22 participants per cluster. However, if the time by cluster effect is stronger, power to demonstrate 5 points change drops to 80%. A 10% increase in sample size will be added to account for non-participation, requiring a total of 30 participants per cluster. Based on the above assumptions, the total minimum sample size is therefore estimated at 30*16=480. The number of participants lost to follow-up will be monitored throughout the study, and mitigation strategies implemented to accommodate any unforeseen impact by additional recruitment of patients. To compute the sample size for each site, a weighting based on the total AYP will be used. The following formula will be used to obtain a sample for each of the study sites.

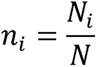

where, *n_i_* = Sample size for site *i, N_i_* = total AYP in site *i and N* = total AYP in all the study sites

### Ethical approval

The AMREF Health Africa Ethics & Scientific Review Committee (ESRC) and the respective County Health Departments have both sanctioned this protocol. Every participant involved provided their informed and written consent. For participants aged between 15 to 17 years, the study investigators were granted a waiver for parental consent by the Ethics and Scientific Review Committee (ESRC). This decision was based on several considerations: to avoid unintentional revelation of the HIV status, to enhance the participation rate, and to mitigate selection bias, especially since the study entailed minimal risk to its participants. Furthermore, the National Commission for Science, Technology, and Innovation (NACOSTI) has provided a research license permitting the execution of the study. Additionally, this trial has been catalogued in the Pan African Clinical Trial Registry at www.pactr.org with the distinctive ID: PACTR202303729957231.

### Dissemination of results

Research results will be disseminated via stakeholder forums, scientific conferences, research publications and the media.

### Study limitations

The experience of end-users of the portal may be affected by loss of smartphones or internet bundles. Application builds may be unstable on various operating system (OS) versions despite round the clock efforts to identify and mitigate these hurdles. Network and power outages may limit access to the portal as well as integration of portal and EMR. These challenges might result in lower-than-anticipated levels of patient activation from the intervention.

### Discussion and Implications

To our knowledge, this is the first study of its kind in Kenya to explore the utility of a smart phone-based application on patient activation among AYP living with HIV. By engaging this tech-savvy sub-population that has been shown to have unique needs as equal partners in their wellness journey through myCareHub®, the study will provide a blue-print that could be applicable to other diseases and sub-populations, more so as features of the intervention itself such as quality, fidelity and usability will be documented.

## Data Availability

This is a protocol and data will be fully available on analysis.

## List of References

1. UNICEF. HIV and AIDS. Protecting children and adolescents from HIV and AIDS and providing care. 2021.

2. Bekker LG, Hosek S. HIV and adolescents: focus on young key populations: J Int AIDS Soc. 2015 Feb 26;18(2Suppl 1). doi: 10.7448/IAS.18.2.20076. eCollection 2015.

3. UNAIDS. AIDSinfo global data on HIV epidemiology and response. 2020.

4. Kemp S. Digital 2021 Kenya2021 14th September 2023. Available from: https://datareportal.com/reports/digital-2021-kenya?rq=kenya.

5. Nations U. YouthStats: Information and communication technology. 2022.

6. Jongbloed K, Parmar S, van der Kop M, Spittal PM, Lester RT. Recent Evidence for Emerging Digital Technologies to Support Global HIV Engagement in Care. Curr HIV/AIDS Rep. 2015;12(4):451–61.

7. Otte-Trojel T, de Bont A, Rundall TG, van de Klundert J. How outcomes are achieved through patient portals: a realist review. Journal of the American Medical Informatics Association : JAMIA. 2014;21(4):751–7.

8. West R, Michie S. A brief introduction to the COM-B Model of behaviour and the PRIME Theory of motivation. Qeios. 2020.

9. WHO. Monitoring and evaluating digital health interventions: a practical guide to conducting research and assessment. Geneva 2016.

10. Marshall R, Beach MC, Saha S, Mori T, Loveless MO, Hibbard JH, et al. Patient activation and improved outcomes in HIV-infected patients. J Gen Intern Med. 2013;28(5):668–74.

11. Hemming K, Kasza J, Hooper R, Forbes A, Taljaard M. A tutorial on sample size calculation for multiple-period cluster randomized parallel, cross-over and stepped-wedge trials using the Shiny CRT Calculator. Int J Epidemiol. 2020;49(3):979–95.

